# Head-to-head comparison demonstrates strong agreement between the harmonized and the self-explanatory versions of the ALS functional rating scale: results of a prospective study

**DOI:** 10.1101/2025.07.21.25331900

**Authors:** André Maier, Yasmin RN Koc, Laura Steinfurth, Dagmar Kettemann, Jenny Norden, Alessio Riitano, Phillip Schmitt, Felix Kolzarek, Senthil Subramanian, Christoph Münch, Susanne Spittel, Thomas Meyer

**Author notes:** Corresponding Author: Dr. André Maier Charité – Universitätsmedizin Berlin Center for ALS and other Motor Neuron Disorders Augustenburger Platz 1, 13353 Berlin, Germany Mail.

## Abstract

**Introduction/Aims:** The ALS Functional Rating Scale-Revised (ALSFRS-R) in its harmonized version is an established outcome parameter in amyotrophic lateral sclerosis (ALS). Assessment of the ALSFRS-R needs standard operating procedures (SOP) for procedural consistency. In contrast, the assessment of the self-explanatory (SE) version of the ALSFRS-R does not include SOPs. In this study, a head-to-head comparison of the harmonized and self-explanatory ALSFRS-R was performed.

**Methods:** In a prospective study, the harmonized ALSFRS-R was assessed in 107 ALS patients, administered by a SOP-trained nurse. In parallel, all patients independently completed the ALSFRS-R-SE, either on a printed form (n = 36) or remotely via the ALS App (n = 71). Agreement between methods was investigated using Spearman’s correlation, Lin’s concordance correlation coefficient (CCC), Deming regression, Bland-Altman plots, and item-level statistics including Kendall’s tau-b and the Stuart-Maxwell test.

**Results:** Total scores from ALSFRS-R and ALSFRS-R-SE showed high correlation and concordance (CCC > 0.9). Deming regression and Bland-Altman analysis revealed no systematic bias. Item-level agreement was generally high, with limited variability in specific items such as handwriting, walking, and dyspnea. ALS progression rates were highly consistent. ALSFRS-R-SE remained robust across remote digital and paper-based assessments.

**Discussion:** The strong agreement observed between the harmonized and self-explanatory versions of the ALSFRS-R supports the interchangeable use of the ALSFRS-R-SE. The self-explanatory format may facilitate remote digital assessment and reduce the complexity of ALSFRS-R assessment in both research and clinical practice. Further studies are warranted to validate the ALSFRS-R-SE across larger cohorts, multiple languages, and diverse rater groups.

## 1 INTRODUCTION

Amyotrophic lateral sclerosis (ALS) is characterized by the progressive loss of motor function. To record motor functions and ALS-related symptoms, the ALS Functional Rating Scale in its revised form (ALSFRS-R), an established instrument in ALS research and clinical care, is used.^1,2^ The scale is administered mainly through an interview conducted by a healthcare professional, with the interviewer undergoing training to ensure that the data is collected as standardized as possible. The standard operating procedures (SOPs) for conducting the interviews have undergone a harmonization process.^3^ Despite the implementation of a harmonized SOP aimed at minimizing inter- and intra-rater variability, fundamental limitations of the standardized ALSFRS-R assessment remain. Thus, the change of evaluators was repeatedly identified as a key factor in sudden changes in the assessment of functional deficits.^4,5^ To overcome the recurring issues of changing evaluators, patient self-assessment has been proposed, leading to the development of several adapted forms of the ALSFRS-R for self-entry.^6–9^ Moreover, a self-explanatory (SE) version of the ALSFRS-R (ALSFRS-R-SE) was developed, which encompasses additional, clearly understandable explanations and specifications for each item.^10^ The ALSFRS-R-SE was consented by an ALS expert group and has been published in English and German. Beyond patient self-rating, the ALSFRS-R-SE is aimed to be used by healthcare professionals, even without training. The ALSFRS-R-SE contributes to the increasing trend towards digitalization and the use of personal smartphones for ALSFRS-R assessment.^11^ Only recently, when the ALSFRS-R-SE was used for remote digital assessment via smartphone, the intrasubject variability, an important confounder in trial settings, was demonstrated to be non-inferior compared to clinical capture.^12^

Despite the broader use of the ALSFRS-R-SE in German-speaking countries, uncertainties persist regarding how the harmonized and self-explanatory versions of the ALSFRS-R may affect the variability.^13^ To address these uncertainties, a head-to-head comparison study was performed in which the harmonized ALSFRS-R, assessed by a SOP-trained study nurse, was compared to the ALSFRS-R-SE, self-administered by the patient, either on a printed form or remotely, using the ALS App.

## 2 METHODS

### 2.1 Study design

This study was a prospective longitudinal single-center study. Conducting and reporting the study was guided by the COnsensus-based Standards for the selection of health Measurement INstruments (COSMIN),^14^ although only part of the complex framework was applied because the measurement properties of the instruments have not been studied in total. The investigation took place between September 2023 and February 2025.

### 2.2 Variables

#### 2.2.1 Amyotrophic Lateral Sclerosis Functional Rating Scale-Revised (ALSFRS-R)

The revised ALS Functional Rating Scale (ALSFRS-R) is a disease-specific, validated instrument for assessing motor impairment in four domains: bulbar, fine motor, gross motor, and respiratory function. It comprises 12 items with 5 scoring options for each item (0 to 4). The total range of the scale is from 0 (no function) to 48 scale points (full function). The four domains are typically analyzed as subscales, as they allow a higher degree of differentiation compared to the total score,^15^ which reflects the overall disease severity.

#### 2.2.2 Harmonized standard operation procedure of administering the ALSFRS-R

The ALSFRS-R and its predecessor, the ALSFRS, were originally designed as a survey instrument for clinical trials, in which healthcare professionals conduct a structured interview with patients about their symptoms and then transcribe the answers into the most appropriate form for the scale items.^1,16^ The trainings that then emerged, first in the USA by NEALS and then also in Europe by TRICALS, pursued the purpose of ensuring that the assessment was as similar as possible in order to reduce intrarater and the far more significant interrater variability.^5^ However, the harmonization between the two entities resulted in an SOP that addresses many of the uncertainties of the original scale and provides some important considerations regarding possible patient responses. With the new SOP, both NEALS and TRICALS are committed to providing training in a consistent manner.^3^

#### 2.2.3 Self-explanatory version of the ALSFRS-R (ALSFRS-R-SE)

The self-explanatory version of the ALSFRS-R was developed in a consensus initiative of a group of German ALS clinicians and researchers.^10^ All participants of the process had extensive experience with the application and evaluation of the ALSFRS-R. The motivation for this activity was the awareness of inconsistent SOPs, the lack of a uniform German version at the time, and the desire to resolve different scales in different settings. The premise of the ALSFRS-R-SE was always to reflect the original scale as closely as possible, without altering it. By providing explanatory notes, easy language, and a clear introductory text, every person, patients and HCPs alike, would be able to perform an ALS assessment without training. This makes the scale a valuable instrument in a clinical and trial environment, where, in addition to traditional assessment situations, the self-evaluation of patients using remote digital methods, such as the ALS app, is becoming increasingly important.^12^

### 2.3 Procedure

#### 2.3.1 Assessment of harmonized ALSFRS-R and ALSRS-R-SE

The aim of this study was to directly compare the ALSFRS-R collected via interview according to the harmonized SOP with the self-documented ALSFRS-R-SE (SE) completed by the participant. The comparison was to be carried out on the same patient using the most common method, in which the respective version of the scale is used. Therefore, the interview was conducted by HCPs and the SE assessment was conducted by the patients themselves. To administer the interview, we selected two experienced ALS study nurses who were trained and certified in the harmonized SOP but had no prior experience working with the ALSFRS-R-SE. The interview assessment was conducted on two consecutive visits in the ALS department. For the SE assessment, independent of the interview, the patient was asked to complete the remote digital assessment using a mobile application (ALS App), which is compatible with both smartphones and tablet devices and is available for iOS and Android devices (https://www.ambulanzpartner.de/als-app/). If patients stated that they had completed the SE on their smartphone a maximum of three days before the visit or that they intended to complete it at home up to three days afterwards, this data was used.

#### 2.3.2 Patients and sample size

The statistical method of choice for presenting the agreement of an assessment instrument that was either collected by different raters or using different methods is to determine the level of concordance using the Bland and Altman method with limits of agreement (LoA),^17^ which allows for a nuanced visualization and quantification of systematic bias and variability between measurements. That formed the basis for the selection of the method of sample size estimation.^18^ In order to calculate the sample size, we used values from preliminary studies that already reported distances of LoA and standard deviations of the differences.^7,8^ In accordance with these studies, the standard deviation of the differences was set to 2.5, and limits of agreement < 7 points were considered acceptable. A value of 0 was chosen for the bias, as the literature reports only minor systematic deviations. Given a significance level of 0.05 and a power of 80%, a total case number of at least 30 subjects per cohort was recommended. Since remote digital assessment was preferred, recruitment at ALS outpatient clinic at Charité continued until the print cohort had reached the projected number.

### 2.4 Protocol approvals and registrations

The study protocol was approved by the Medical Ethics Committee of Charité - Universitätsmedizin Berlin, Germany (EA2/190/23). Written informed consent was obtained from all participants.

### 2.5 Statistical methods

Descriptive statistics were used for the statistical analysis (mean, standard deviation inLJ±LJand ranges). The selection of methods for comparing the instruments, the harmonized ALSFRS-R interview and ALSFRS-R-SE, followed the Recommendations of COSMIN and the ISPOR ePRO Task Force.^19^ As an interview-based questionnaire, it was compared with a paper-based and an electronic PRO (ePRO) that represents a further development of the questionnaire. To assess the relationship and agreement between the two versions of the questionnaire in two cohorts, multiple statistical approaches were applied. Spearman’s rank correlation coefficient was used to evaluate the monotonic association between the total scores of the ALSFRS-R interview and ALSFRS-R-SE. In addition, Lin’s Concordance Correlation Coefficient (CCC) was calculated to quantify the concordance by accounting for both precision and accuracy.^20^ To further examine potential systematic differences and proportional bias between the two score sets, a Deming regression was conducted.^21^ This regression method accounts for measurement error in both variables by estimating a line of best fit that minimizes the orthogonal distances between the observed data points and the regression line. The resulting intercept and slope parameters were interpreted to assess fixed and proportional bias, respectively. Additionally, a Bland-Altman analysis was performed to visualize the agreement between the two measurement methods and to identify any systematic bias.^17^ Mean differences and 95% limits of agreement (mean ± 1.96 SD) were calculated and plotted against the mean of the two measurements. To evaluate the agreement and consistency of individual item responses, the following statistical methods were employed.

Mean differences between item scores were calculated to assess potential systematic shifts in response tendencies across modalities. Kendall’s Tau-b was used to evaluate the ordinal association between item responses, providing a non-parametric measure of rank correlation that accounts for ties.^22^ This was particularly used given the ordinal nature of the item response scales. To assess marginal homogeneity across paired categorical responses, the Stuart-Maxwell test was applied.^23^ This test examines whether the distribution of item responses differs significantly between the two administration modes, allowing the detection of systematic shifts in response patterns at the item level. All analyses were conducted using R version 4.4.2 (2024-10-31), and statistical significance was set at p < 0.05.

## 3 Results

### 3.1 Demographic and clinical characteristics

This study included 107 ALS patients who provided at least one self-assessed ALSFRS-R-SE (SE) data set, either on the App or on a printed form, and a harmonized ALSFRS-R interview. Follow-up visits were carried out on 81 patients after 5.4 months on average (SD 55.5) of the initial visit (**Figure 1**).

**Figure 1:**
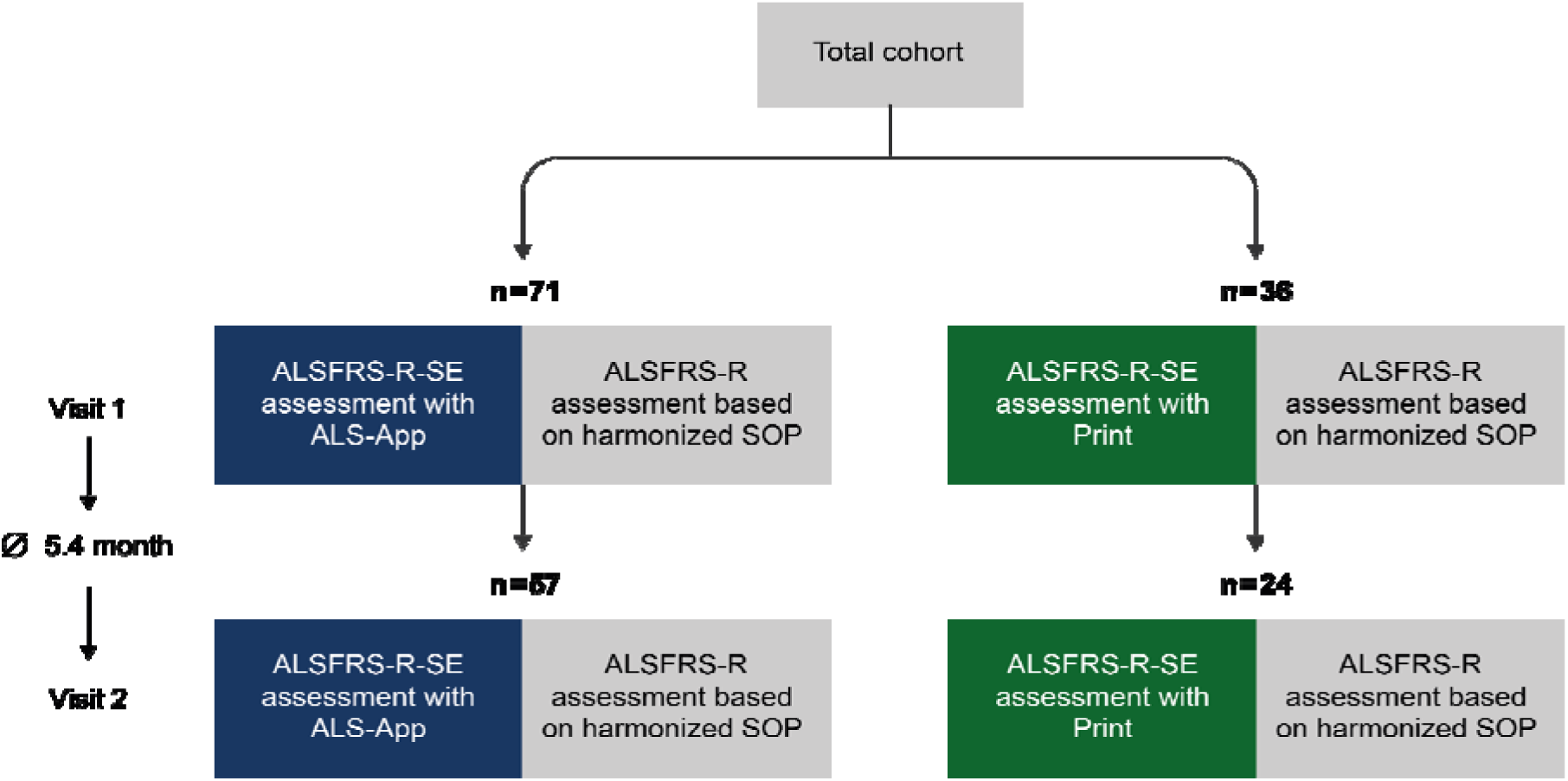
Flow chart of the patient cohort. Participants were assigned to one of two groups depending on the mode of ALSFRS-R-SE collection: remote digital assessment via the ALS App or completion on a printed form. At two study visits, the ALSFRS-R was administered through an interview based on the harmonized SOP.

The demographics and clinical characteristics of the participants are presented in **Table 1**. There were no differences between the ALS App cohort and the print cohort with respect to sex distribution. Participants who completed the ALSFRS-R-SE on paper had significantly higher values for age, disease duration, and progression rate. There was a non-significant trend toward greater disease severity for these participants, as indicated by lower ALSFRS-R scores.

**Table 1:** Demographic and clinical characteristics of participants at baseline. n=107

| Characteristics | Classification | Total cohort | ALS App cohort | Print cohort | p-value<sup>1</sup> |
| --- | --- | --- | --- | --- | --- |
| Sex | female, % (n) | 39.4 (43) | 39.4 (28) | 41.7 (15) | 0.824 |
|  | male, % (n) | 60.6 (64) | 60.6 (43) | 58.3 (21) |  |
| Age | at onset, years, mean (SD, R) | 58.4 (10.6, 25.4- 83.9) | 57.7 (10.2; 25.4-83.9) | 59.8 (11.3; 30.6-81.5) | 0.272 |
|  | at visit 1, years, mean (SD, R) | 63.2 (9.4, 41.7-87.2) | 61.2 (8.8; 41.7-84.9) | 67.2 (9.4; 44.2-87.2) | 0.001 |
| Disease duration | at visit 1, months, median (IQR) | 29.77 (15.9-55.5) | 28.9 (14-45) | 40.3 (20.6-141.1) | 0.003 |
| ALSFRS-R-SE total score (max. 48) | mean (SD, R) | 35.1 (8.3, 10-47) | 35.6 (8.1; 9-47) | 33.4 (8.5; 11-46) | 0.059 |
| ALSFRS-R interview (max. 48) | mean (SD, R) | 34.9 (8.3, 9-47) | 36.1 (8.1; 12-47) | 33.1 (8.4; 10-46) | 0.156 |
| Rate of disease progression | mean (SD, R) | 0.50 (0.54, 0.01, 3.70) | 0.57 (0.4; 0.01-3.70) | 0.37 (0.04-2.79) | 0.049 |

Abbreviations: n, number of participants; SD, standard deviation; R, range; ALSFRS-R-SE, Amyotrophic Lateral Sclerosis Functional Rating Scale, revised, self-explanatory; ALSFRS-R, Amyotrophic Lateral Sclerosis Functional Rating Scale, revised; IQR, interquartile range. 1 Pearson's Chi-squared test; Wilcoxon rank sum test

### 3.2 Comparison of harmonized ALSFRS-R and ALSFRS-R-SE total scores

#### 3.2.1 Correlation, Concordance and Regression of harmonized ALSFRS-R and ALSFRS-R-SE total scores

Various statistical methods were used to compare the total values, demonstrating not only the level of agreement but also the degree of deviation between them. In all visits, both in the ALS App and in the print cohort, there was a high Spearman’s correlation and concordance (CCC) between the total values with coefficients above 0.9 (**Table 2**). The Deming regression revealed no statistically significant difference from the estimated line of best fit, as no fixed or proportional bias was found, neither at the individual nor at the aggregated time points (**Figure 2**; **Table 2**).

**Figure 2:**
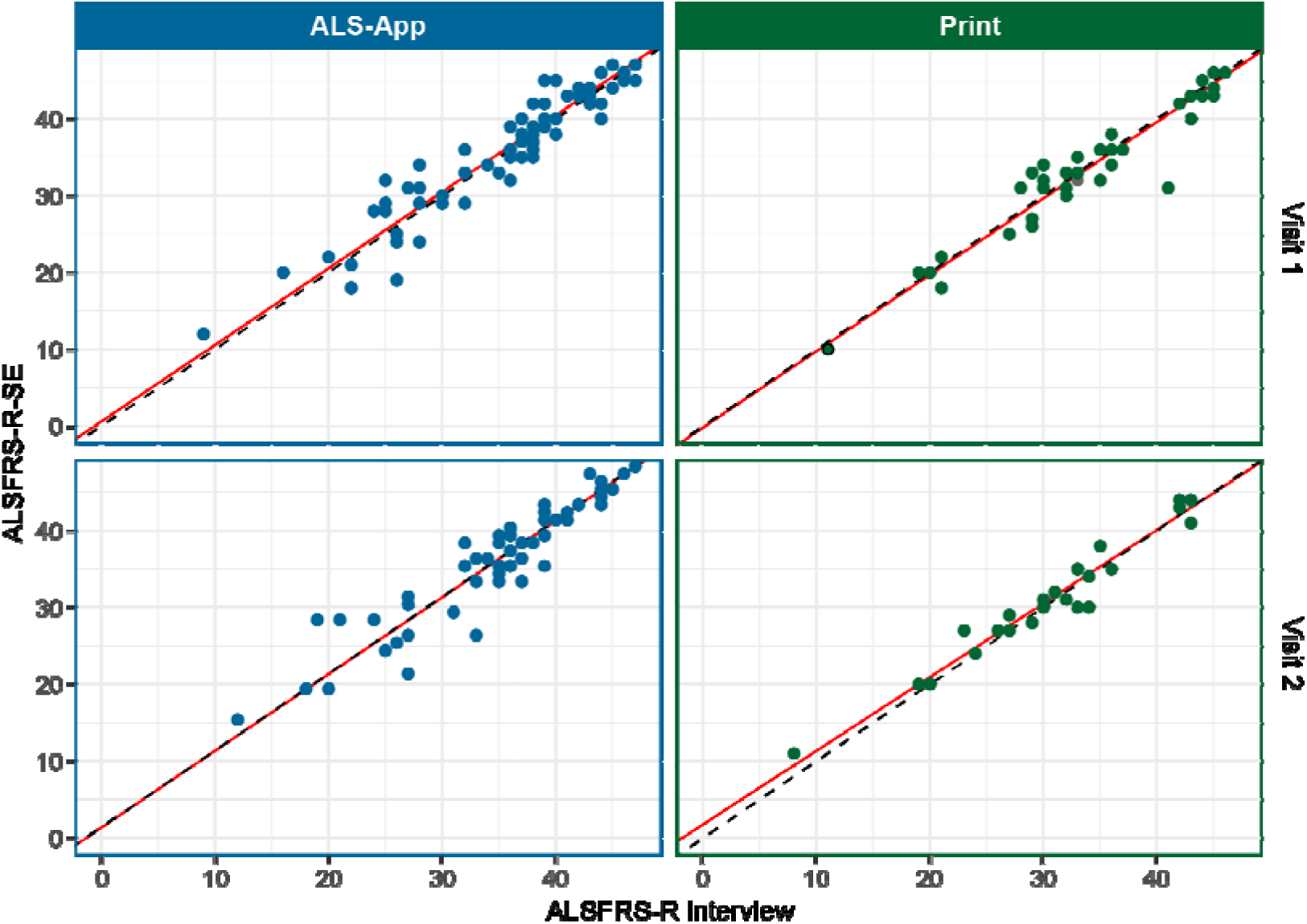
Correlation and Deming-Regression of harmonized ALSFRS-R interview and ALSFRS-R-SE total scores across the two cohorts of ALS App and paper based ALSFRS-R-SE assessment and at two timepoints (visit 1 and 2). The correlation of the total scores is shown by the proximity to the bisecting line (dashed line). Deming regression is represented by the deviation (intercept and slope) of the bisecting line and the estimated line of best fit (red line).

**Table 2:** Correlation, concordance and regression of total scores between the harmonized ALSFRS-R SOP, which was recorded as an interview, and the ALSFRS-R SE, which was completed by the patient at two time points. The SE was documented either in the ALS app or on a printed form.

| Cohort | Visit | N | Correlation |  | Concordance |  | Regression<sup>2</sup> |  |  |  |  |  |
| --- | --- | --- | --- | --- | --- | --- | --- | --- | --- | --- | --- | --- |
|  |  |  | Spearman's | p-value | CCC<sup>1</sup> | 95% CI | Intercept | 95% CI | p-value | Slope | 95% CI | p-value |
| App | 1 | 71 | 0.95 | <0.001 | 0.97 | 0.91; 0.97 | 0.6 | -3.17; 4.37 | 0.76 | 1 | 0.9; 1.09 | 0.95 |
| App | 2 | 57 | 0.92 | <0.001 | 0.94 | 0.88; 0.96 | -0.04 | -4.8; 4.71 | 0.99 | 1 | 0.88; 1.12 | 0.98 |
| Print | 1 | 36 | 0.91 | <0.001 | 0.98 | 0.91; 0.98 | -0.16 | -2.52; 2.19 | 0.89 | 0.99 | 0.93; 1.06 | 0.86 |
| Print | 2 | 24 | 0.95 | <0.001 | 0.97 | 0.93; 0.99 | 1.77 | -1.39; 4.93 | 0.26 | 0.96 | 0.86; 1.06 | 0.41 |

<sup>1</sup>Concordance Correlation Coefficient; <sup>2</sup>Deaming Regression

#### 3.2.2 Systemic Error and Agreement of harmonized ALSFRS-R and ALSFRS-R-SE total scores

The systematic error is represented by the mean difference between the mean total scores. There was no statistically significant difference or specific deviation in one direction. Patients in the largest group, who rated themselves via the ALS App at visit 1, tended to rate themselves slightly higher. However, this was not the case for visit 2 or in the print cohort visit 1, but again in the smallest group of the print cohort at visit 2 (**Table 3**). The coefficients of variation (CoV), i.e., the relative standard deviation, are also nearly equal, which reflects a comparable distribution in the two versions of ALSFRS-R.

**Table 3:** Systemic Error and Agreement of total scores between the harmonized ALSFRS-R SOP, which was recorded as an interview, and the ALSFRS-R SE, which was completed by the patient at two time points. The SE was documented either in the ALS app or on a printed form.

| Cohort | Visit | N | Harmonized ALSFRS-R |  | ALSFRS-R-SE |  | Difference |  | Limits of Agreement |  |  |
| --- | --- | --- | --- | --- | --- | --- | --- | --- | --- | --- | --- |
|  |  |  | Mean (SD) | CoV<sup>1</sup> | Mean (SD) | CoV<sup>1</sup> | Mean (SD) | p-value<sup>2</sup> | Lower Limit | Upper Limit | 95% CI |
| ALS App | 1 | 71 | 35.63 (8.12) | 0.23 | 36.13 (8.09) | 0.22 | -0.49 (2.64) | 0.11 | -5.67 | 4.68 | -6.66; 5.67 |
| ALS App | 2 | 57 | 34.79 (7.58) | 0.22 | 34.7 (7.57) | 0.22 | 0.09 (2.83) | 1 | -5.46 | 5.63 | -6.66; 6.84 |
| Print | 1 | 36 | 33.44 (8.47) | 0.25 | 33.08 (8.42) | 0.25 | 0.36 (2.5) | 0.47 | -4.53 | 5.26 | -4.53; 6.65 |
| Print | 2 | 24 | 30.58 (8.26) | 0.27 | 31.08 (7.92) | 0.27 | -0.5 (1.89) | 0.18 | -4.2 | 3.2 | -5.56; 4.56 |

1 Coefficient of Variation; 2 Wilcoxon rank sum test

The agreement between the harmonized ALSFRS-R and ALSFRS-R-SE was examined using the Bland-Altman method. This showed that the agreement between the ALSFRS-R version was very high in both cohorts and at both timepoints at visits 1 and 2 (**Figure 3**, **Table 3**). More than 95% of all pairs of measurements were within the limits of agreement (LoA). Except for visit 2 in the print cohort, the LoA was comparable across all conditions. The narrower range suggests a higher level of agreement; however, the limited number of cases warrants caution when interpreting this finding.

**Figure 3:**
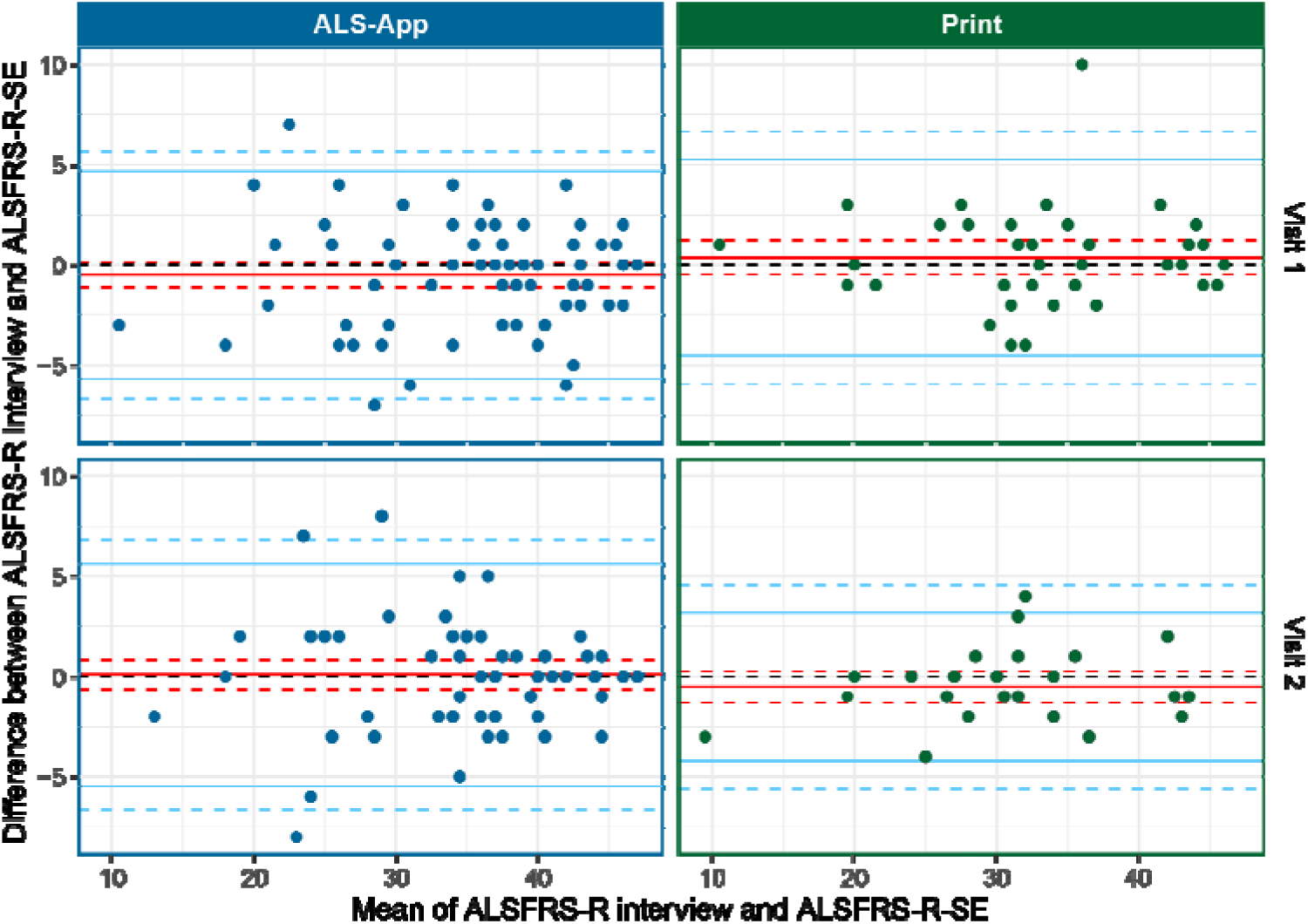
Bland-Altman-Plot of harmonized ALSFRS-R interview and ALSFRS-R-SE total scores across the two cohorts of App and paper based ALSFRS-R-SE assessment and at two timepoints (visit 1 and 2). The line of mean differences (bias) is shown as a continuous red line, while a dashed red line indicates the 95% confidence intervals. The limits of agreement are shown as a continuous blue line, with a dashed blue line representing its 95% confidence interval.

### 3.3 Single-item analysis of harmonized ALSFRS-R and ALSFRS-R-SE

As the individual items may behave differently compared to the overall score or each other, an analysis of the single items was carried out. The absolute difference between the individual items is on average only 0.12 (SD 0.1; range - 0.25-0.21) (**Supplement Table S1**).

The agreement of the individual items and measures for assessing ordinal characteristics (Kendall’s tau and Spearman’s correlation) was used to determine how the items in the two versions of the ALSFRS-R resemble each other. The study revealed that although the agreement of most items was high, certain items showed only moderate agreement values in individual visits (**Table 4**). Uncertainty intervals that reached below 50% were observed for item 4 (handwriting; in the print cohort at visit 1 and 2), item 5 (cutting food/handling utensils; in the print cohort at visit 2), item 8 (walking; in the print cohort at visit 2), item 9 (climbing stairs; in the app cohort at visit 1 and print cohort at visit 2), and item 10 (dyspnea, in the print cohort at visit 1 and 2). However, it is noticeable that the rank correlation, as assessed by Kendall’s τ coefficient, remains high for these items. In contrast, some items exhibit low rank correlations (e.g., item 11 in the app cohort at visit 1, or item 10 in the print cohort at visit 2), resulting in good percent agreement. This occurs when a large number of measurement pairs are identical; however, if there are deviations, those are unsystematic (**Figure 4**). In accordance with this, the Stuart-Maxwell test consistently shows that there were no systematic deviations in these pairs, as in all others. These unsystematic deviations can also be seen graphically (**Supplement Figure S1**), with an average percentage of disagreement being 11.7% (SD 6.0, range 0.0-29.2) in either direction (**Supplement Table S1**). However, no item in particular stands out as exhibiting particularly low levels of agreement across both cohorts and visits.

**Figure 4:**
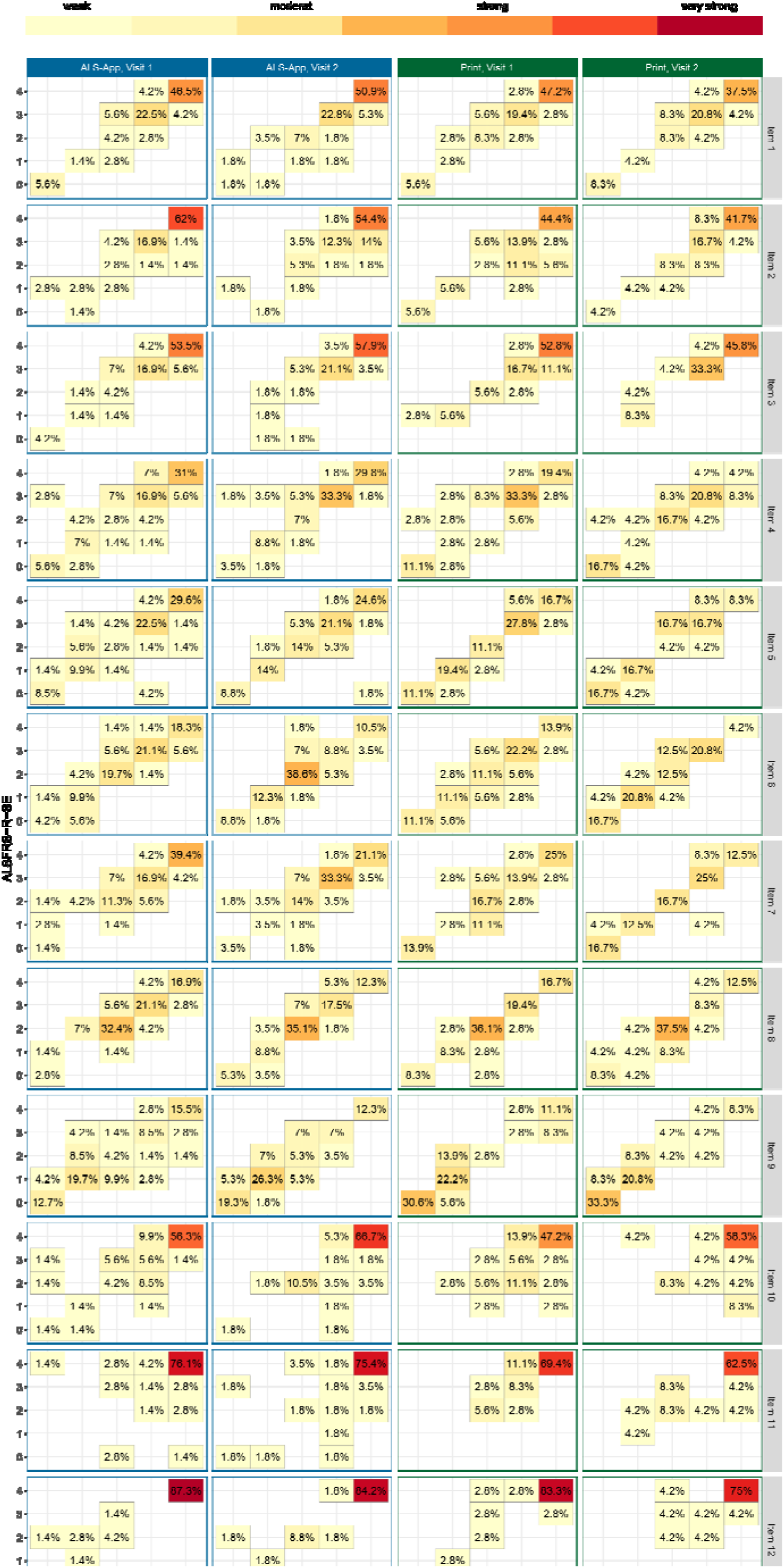
Single-item analysis of the comparison between harmonized ALSFRS-R interview (x-axis) and ALSFRS-R-SE (y-axis). The proportion of patients with the corresponding characteristics of the respective ALSFRS-R versions is shown in each field. A darker color indicates a higher proportion with a stronger correlation, respectively. Fields on the bisecting axis indicate an exact match.

**Table 4:**
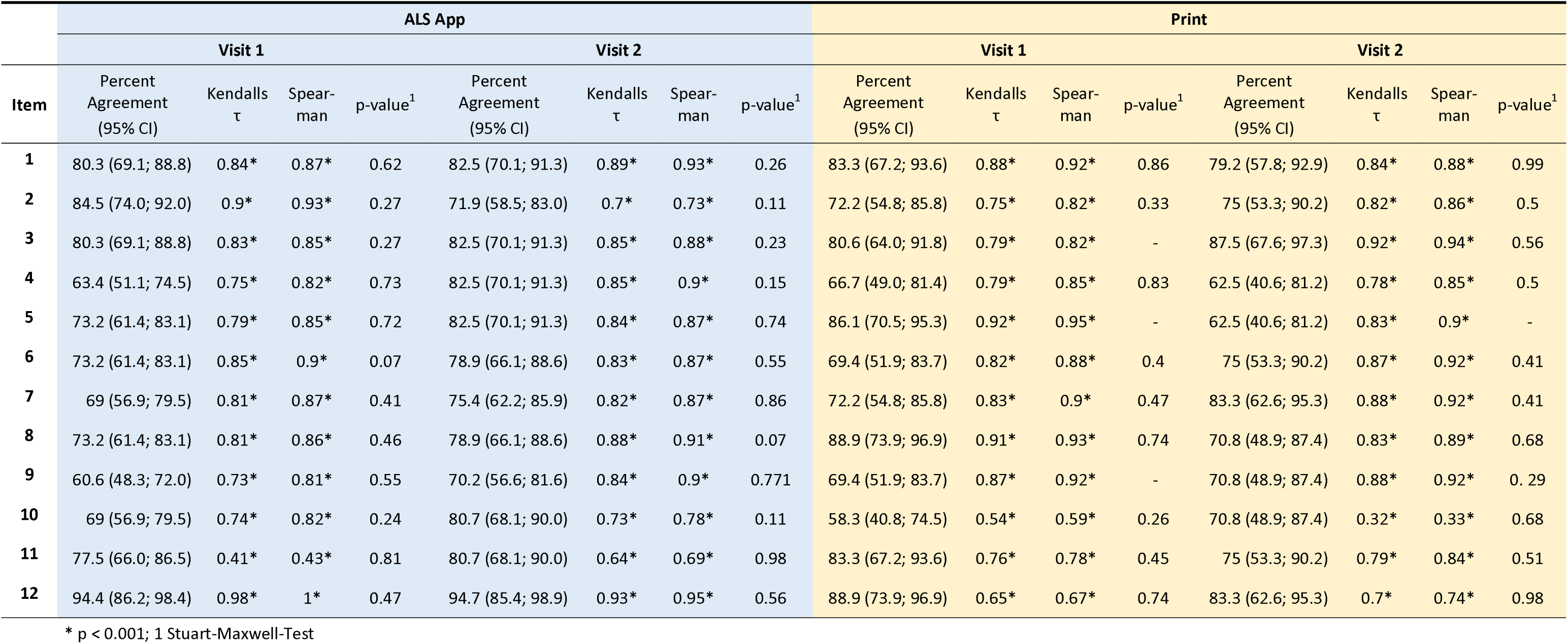
Agreement and correlation of individual ALSFRS-R items between the harmonized ALSFRS-R SOP recorded as an interview and the ALSFRS-R-SE completed by the patient at two timepoints. The SE was documented either in the ALS app or on paper. For empty table cells, the Stuart-Maxwell test could not be applied due to low variability.

### 3.4 Comparison of progression rates

The progression rates (PR) were determined because changes in PR often serve as study outcomes, and differences in PR may impact the statistical analysis.^12^ Subsequently, a categorization of the progression rates into slow, medium, and fast was performed according to Kimura et al.^24^ (**Supplement Table S2**). The categorical evaluation revealed high agreement values **(Supplement Table S3**) and a high correlation in all categories (**Supplement Figure S2**). There was no relevant or statistically significant systematic deviation between the PR of the two ALSFRS-R versions of the harmonized SOP interview and the ALSFRS-R-SE.

## 4 Discussion

### 4.1 Association

Through statistical association and concordance analysis, this study demonstrates a high degree of correspondence between the total values obtained from both methods. A further regression analysis did not reveal any significant deviation from the line of best fit. Therefore, no bias was detected between the ALSFRS-R obtained via interview and the ALSFRS-R-SE documented by the patients.

### 4.2 Agreement

The agreement of ALSFRS-R total values analyzed using the method by Bland and Altman was significant in both groups and at both time points, consistent with the known and expected high variability of total ALSFRS-R values. In contrast to some previous reports,^9,25^ there was no evidence of a consistent systematic bias, suggesting that patients tend to provide generally higher ratings when using self-assessment. The underlying reasons remain speculative. However, it is conceivable that the ALSFRS-R-SE, with its unambiguous phrasing, enables patients to provide assessments more closely aligned with those of interviewers. Moreover, the reduced complexity and enhanced clarity of the ALSFRS-R-SE may contribute to its broader applicability among HCPs who are not highly specialized in ALS. This broader usability can facilitate implementation in more diverse clinical settings and support longitudinal monitoring outside of expert centers. In addition, the intuitive structure of the ALSFRS-R-SE substantially reduces the need for extensive rater training, which in turn may lower training-related costs in clinical trials and registries. This aspect is particularly relevant given the increasing demand for scalable and cost-efficient assessment tools in multicenter studies.

### 4.3 Comparison of the single ALSFRS-R items

This study showed that, overall, the agreement across individual items was high. However, there were individual items that showed a higher deviation, although this was not systematic in one particular direction and rank correlation was still high. For example, the items “climbing stairs” (item 9) and “dyspnea” (item 10) have a reduced percentage agreement with a relatively wide confidence interval. It can be assumed that both the trained HCP and the patient themselves have the most difficulty providing consistent answers based on what has been determined by either the SOP plus training or the explanatory language inside the ALSFRS-R-SE. These differences at the level of individual items also reinforce the notion that the ALSFRS-R should not be interpreted as a total score, but rather should be analyzed in a more differentiated approach, for example, by considering the subscales.^26–28^

### 4.4 Study Limitations

The primary limitations of this study are its limited scale and monocentric design. A larger patient cohort and the involvement of additional trained interviewers could potentially have increased the reliability and robustness of the data. Implementation on a broader scale, such as a parallel project within a large multicenter clinical trial, was not feasible due to resource constraints. Nonetheless, the study included patients from a real-world clinical setting, some of whom had a long disease duration or advanced functional impairment. Participation in the ALS app–based assessment was voluntary, which reflects realistic usage conditions. The reduced number of follow-up visits observed in this context is consistent with such settings. Another limitation concerns the generalizability of the findings to other settings where ALSFRS-R data are collected. In this study, assessments were conducted by experienced interviewers trained according to the established SOP,^3^ and some patients may have been familiar with the ALSFRS-R-SE due to its routine use at the study center.^12^ These conditions, both in terms of interviewer training and patient familiarity, cannot be assumed in other contexts, potentially introducing greater variability, especially in clinical trials. Moreover, cultural or linguistic differences may further influence the reliability and comparability of ALSFRS-R-SE assessments in broader applications.

### 4.5 Conclusion

This study presents a head-to-head comparison between the conventional, interviewer-administered ALSFRS-R and the self-administered ALSFRS-R-SE. The results demonstrate a high level of agreement between both methods. Importantly, the observed variability cannot be attributed to either approach, as no relevant systematic differences in score distribution were found. A particular strength of this work lies in the item-level analysis, which revealed no consistent directional bias. Nonetheless, individual items showed differing patterns, highlighting the value of further item-specific investigation. The ALSFRS-R-SE has already been successfully implemented as an electronic patient-reported outcome (ePRO) within the ALS app, based on a Bring Your Own Device (BYOD) model.^29^ The wide availability of the tool in this digital format is contributing to ongoing data collection in real-world settings, and further datasets from this application are expected. The added value of remote assessment is also underscored by recent data showing that over three quarters of patients consider the functional rating scale to be prognostically relevant, while fewer than 10% experience the self-assessment as burdensome.^30^

Importantly, the present results are in line with findings from a previous non-inferiority study, in which a subset of participants had already used the ALSFRS-R-SE.^12^ This connection between the two study cohorts not only lends further support to the robustness of the current findings but also confirms its reliability as an alternative to interview-based administration. Beyond its clinical usability, the ALSFRS-R-SE has also been adopted in a large-scale multicenter biomarker study, where it serves as the primary functional outcome measure.^31^ This will enable a direct comparison between ALSFRS-R-SE trajectories and the longitudinal dynamics of neurofilament light chain (NfL), providing important insights into disease progression. Furthermore, future studies are planned to explore the integration of ALSFRS-R-SE in clinical routines involving a broader range of healthcare professionals, thereby supporting its continued validation across different user groups.

Looking ahead, it will be essential to ensure that the ALSFRS-R-SE is adaptable across cultural and linguistic contexts. In such settings, acceptance of the instrument may be achieved without repeated formal validation, as long as expert involvement ensures conceptual and contextual consistency.^32^ This perspective aligns with the recent call for harmonization of self-administered ALSFRS-R, which highlighted the variability in item phrasing and structure across existing versions as a major obstacle to international comparability.^13^ In this context, the consented ALSFRS-R-SE may serve not only as a practical alternative to interview-based assessments, but as a mid-term candidate for replacing the current version of the ALSFRS-R. Achieving such a transition, however, will require robust validation that supports regulatory acceptance by authorities such as the EMA and FDA. The present study contributes critical initial evidence for the ALSFRSl7lRl7lSE in the German language, which is particularly relevant given that Germanl7lspeaking countries represent a substantial share of ALS patients within the EMA regulatory framework.

## Data Availability

All data produced in the present study are available upon reasonable request to the authors.

## Author Contributions

**André Maier:** conceptualization, writing – original draft, methodology, writing – review and editing, investigation, formal analysis, data curation, project administration. **Yasmin Koc:** investigation, data curation. **Laura Steinfurth:** conceptualization, investigation, data analysis. **Dagmar Kettemann:** writing – review and editing, investigation. **Jenny Norden:** writing – review and editing, investigation. **Alessio Riitano:** investigation, data curation. **Phillip Schmitt: Felix Kolzarek:** investigation, data curation. **Senthil Subramanian:** writing – review and editing, data analysis. **Christoph Münch:** supervision, resources, data curation. **Susanne Spittel:** writing – review and editing, data analysis. **Thomas Meyer:** writing – review and editing, project administration, supervision, resources, data curation, conceptualization, funding acquisition.

## Acknowledgments

We gratefully acknowledge the participants and their caregivers for their valuable contributions to this study. We also thank the supporting organizations and institutions for enabling and facilitating this work.

## Funding information

This work was supported by the Boris Canessa ALS Stiftung (Düsseldorf, Germany) and Martin Herrenknecht Fonds for ALS Research (H4017703513237604).

## Conflicts of interest statement

AM is on an advisory board of Novartis and has received consulting fees and honoraria for presentations from Zambon GmbH, ITF Pharma, and Roche. TM is on the advisory board of Biogen and has received consulting fees from Biogen. TM and CM are founders and shareholders of the Ambulanzpartner Soziotechnologie APST GmbH, which makes the internet platform Ambulanzpartner and the mobile application “ALS App.” The remaining authors have no conflicts of interest.

## Data availability statement

The data that support the findings of this study are available from the corresponding author upon reasonable request.

## List of abbreviations

ALS: amyotrophic lateral sclerosis
ALSFRS-R: ALS Functional Rating Scale – Revised
ALSFRS-R-SE: ALS Functional Rating Scale – Revised – self-explanatory
CCC: concordance correlation coefficient
CoV: Coefficient of Variation
HCP: health care professional
LoA: Limits of Agreement
PR: progression rate
SOP: standard operating procedure

## Supplement material

**Supplement Table S1:** Mean difference and percentage deviation upward and downward of individual ALSFRS-R items between the harmonized ALSFRS-R SOP, which was recorded as an interview, and the ALSFRS-R SE, which was completed by the patient at two time points. The SE was documented either in the ALS app or on a printed form.

| Item | ALS App |  |  |  |  |  | Print |  |  |  |  |  |
| --- | --- | --- | --- | --- | --- | --- | --- | --- | --- | --- | --- | --- |
|  | Visit 1 |  |  | Visit 2 |  |  | Visit 1 |  |  | Visit 2 |  |  |
| | $\Delta M$ | Percent higher<br>(95% CI) | Percent lower<br>(95% CI) | $\Delta M$ | Percent higher<br>(95% CI) | Percent lower<br>(95% CI) | $\Delta M$ | Percent higher<br>(95% CI) | Percent lower<br>(95% CI) | $\Delta M$ | Percent higher<br>(95% CI) | Percent lower<br>(95% CI) |
| 1 | 0.00 | 9.9 (4.1; 19.3) | 9.9 (4.1; 19.3) | -0.09 | 5.3 (1.1; 14.6) | 12.3 (5.1; 23.7) | 0.06 | 11.1 (3.1; 26.1) | 5.6 (0.7; 18.7) | 0.04 | 12.5 (2.7; 32.4) | 8.3 (1; 27) |
| 2 | -0.03 | 7 (2.3; 15.7) | 8.5 (3.2; 17.5) | -0.16 | 7 (1.9; 17) | 21.1 (11.4; 33.9) | -0.25 | 5.6 (0.7; 18.7) | 22.2 (10.1; 39.2) | -0.08 | 8.3 (1; 27) | 16.7 (4.7; 37.4) |
| 3 | 0.06 | 12.7 (6; 22.7) | 7 (2.3; 15.7) | 0.02 | 10.5 (4; 21.5) | 7 (1.9; 17) | -0.08 | 5.6 (0.7; 18.7) | 13.9 (4.7; 29.5) | 0.12 | 12.5 (2.7; 32.4) | 0 (0; 14.2) |
| 4 | 0.10 | 21.1 (12.3; 32.4) | 15.5 (8; 26) | 0.14 | 12.3 (5.1; 23.7) | 5.3 (1.1; 14.6) | 0.11 | 19.4 (8.2; 36) | 13.9 (4.7; 29.5) | 0.08 | 20.8 (7.1; 42.2) | 16.7 (4.7; 37.4) |
| 5 | -0.01 | 16.9 (9; 27.7) | 9.9 (4.1; 19.3) | -0.05 | 8.8 (2.9; 19.3) | 8.8 (2.9; 19.3) | -0.03 | 5.6 (0.7; 18.7) | 8.3 (1.8; 22.5) | 0.21 | 29.2 (12.6; 51.1) | 8.3 (1; 27) |
| 6 | 0.03 | 14.1 (7; 24.4) | 12.7 (6; 22.7) | -0.02 | 8.8 (2.9; 19.3) | 12.3 (5.1; 23.7) | -0.17 | 8.3 (1.8; 22.5) | 22.2 (10.1; 39.2) | 0.17 | 20.8 (7.1; 42.2) | 4.2 (0.1; 21.1) |
| 7 | 0.10 | 19.7 (11.2; 30.9) | 11.3 (5; 21) | 0.04 | 14 (6.3; 25.8) | 10.5 (4; 21.5) | -0.03 | 11.1 (3.1; 26.1) | 16.7 (6.4; 32.8) | 0.04 | 12.5 (2.7; 32.4) | 4.2 (0.1; 21.1) |
| 8 | 0.10 | 18.3 (10.1; 29.3) | 8.5 (3.2; 17.5) | 0.11 | 15.8 (7.5; 27.9) | 5.3 (1.1; 14.6) | -0.08 | 2.8 (0.1; 14.5) | 8.3 (1.8; 22.5) | -0.04 | 12.5 (2.7; 32.4) | 16.7 (4.7; 37.4) |
| 9 | 0.03 | 21.1 (12.3; 32.4) | 18.3 (10.1; 29.3) | 0.09 | 19.3 (10; 31.9) | 10.5 (4; 21.5) | 0.03 | 16.7 (6.4; 32.8) | 13.9 (4.7; 29.5) | 0.21 | 25 (9.8; 46.7) | 4.2 (0.1; 21.1) |
| 10 | 0.08 | 18.3 (10.1; 29.3) | 12.7 (6; 22.7) | -0.14 | 7 (1.9; 17) | 12.3 (5.1; 23.7) | -0.11 | 19.4 (8.2; 36) | 22.2 (10.1; 39.2) | -0.25 | 8.3 (1; 27) | 20.8 (7.1; 42.2) |
| 11 | -0.03 | 11.3 (5; 21) | 11.3 (5; 21) | -0.05 | 7 (1.9; 17) | 12.3 (5.1; 23.7) | 0.11 | 13.9 (4.7; 29.5) | 2.8 (0.1; 14.5) | -0.04 | 12.5 (2.7; 32.4) | 12.5 (2.7; 32.4) |
| 12 | 0.07 | 5.6 (1.6; 13.8) | 0 (0; 5.1) | 0.04 | 3.5 (0.4; 12.1) | 1.8 (0; 9.4) | 0.08 | 8.3 (1.8; 22.5) | 2.8 (0.1; 14.5) | 0.04 | 8.3 (1; 27) | 8.3 (1; 27) |

$\Delta M$ = mean difference

**Supplement Figure S1:**
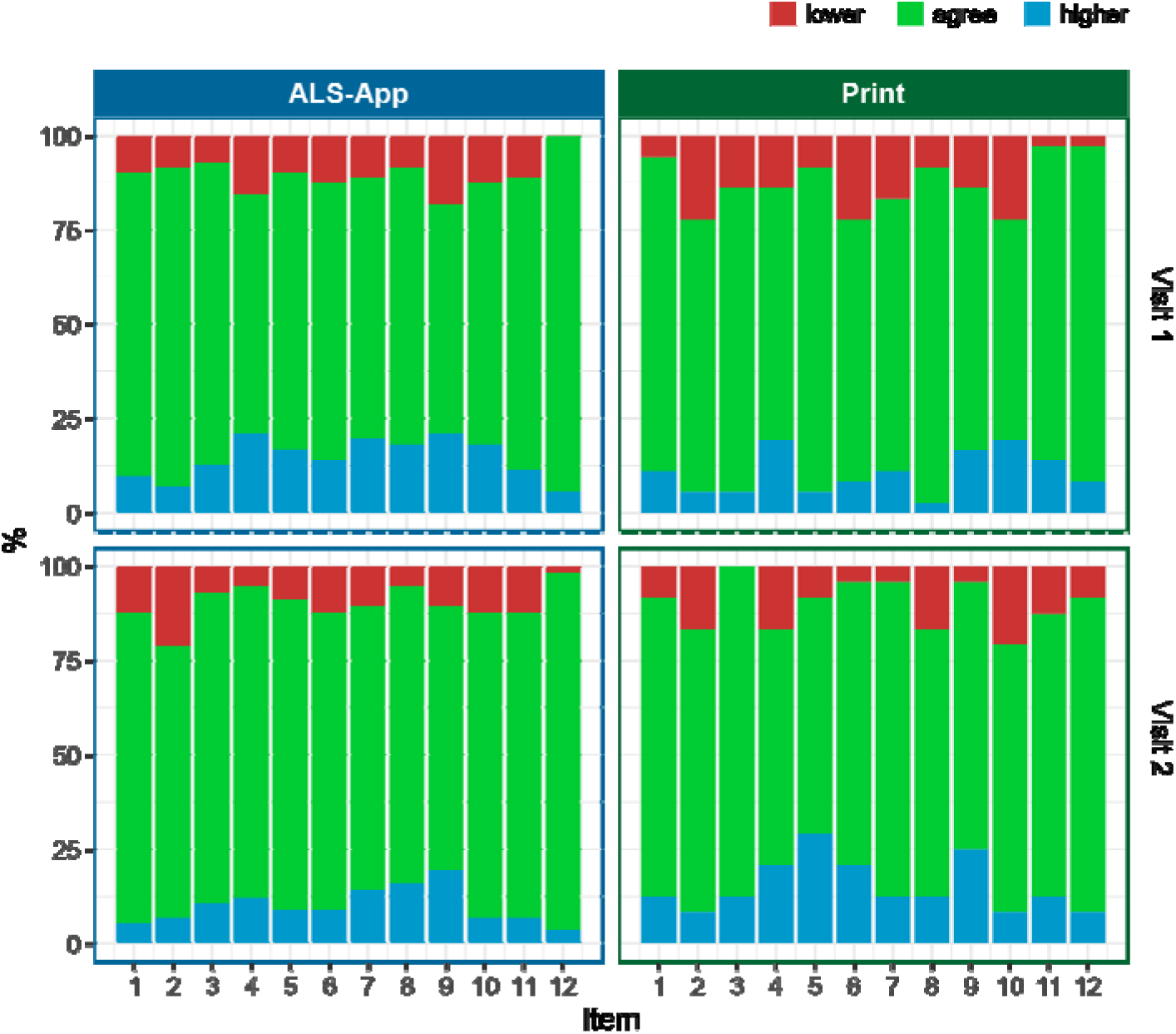
Proportion in percent of agreement and deviation of the singe items of harmonized ALSFRS-R interview and ALSFRS-R-SE. The agreement between the measurements is represented by green color, the deviation upwards by red and downwards by blue.

**Supplement Table S2:**
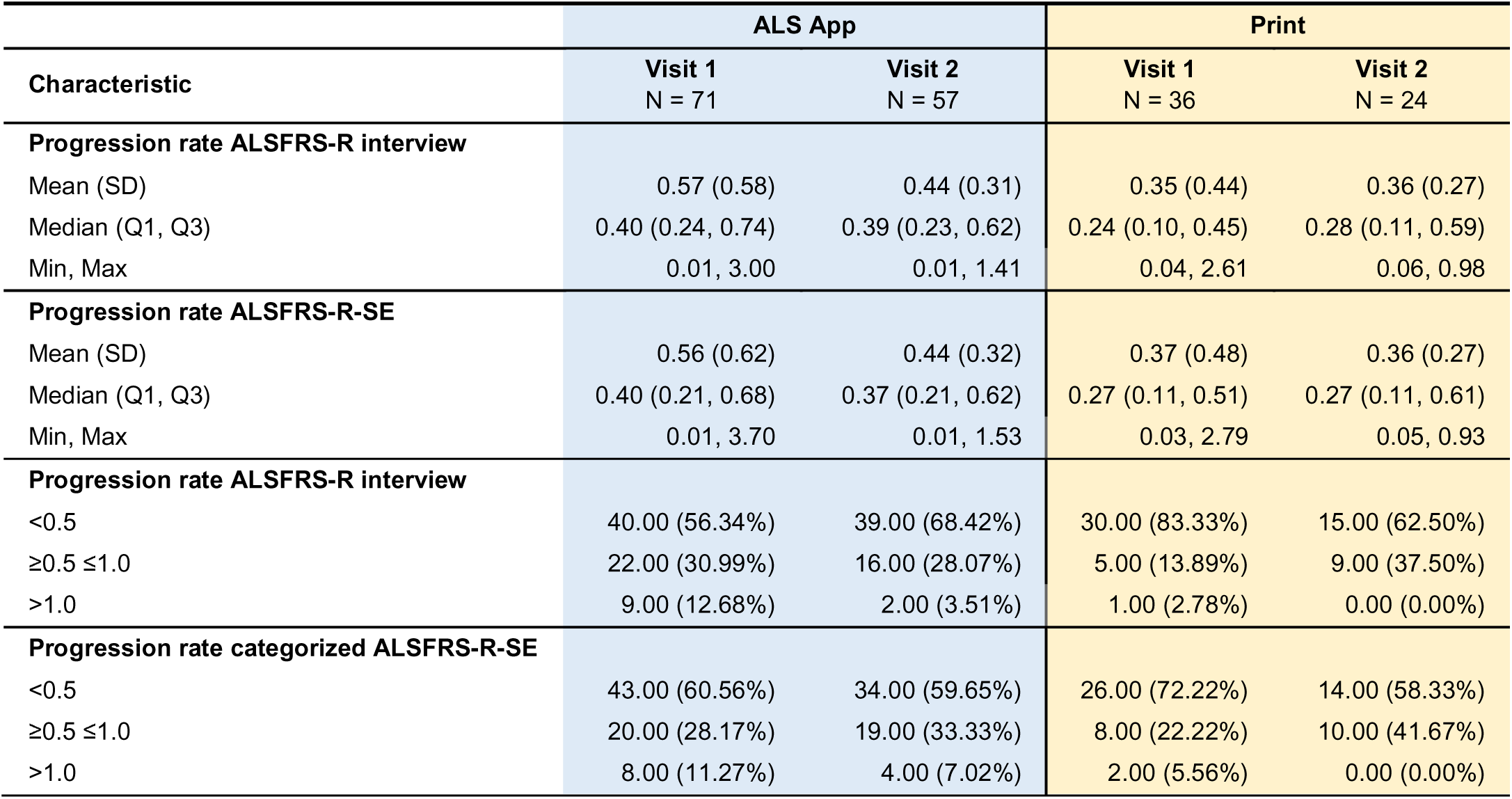
Mean progression rate in the individual cohorts and across visits. Categorization of PR was carried out in <0.5, ≥ 0.5 to ≤ 1.0, and greater than 1.0. The distribution of patients is reported in the categories.

**Supplement Table S3:** Agreement and correlation of progression rates between the harmonized ALSFRS-R SOP recorded as an interview and the ALSFRS-R-SE completed by the patient at two timepoints.

| Kohort | Visit | Percent Agreement<br>(95% CI) | Kendalls $\tau$ | Spearman | p-value<sup>1</sup> |
| --- | --- | --- | --- | --- | --- |
| ALS App | 1 | 94.4 (86.2; 98.4) | 0.93* | 0.94 | 0.135 |
|  | 2 | 80.7 (68.1; 90.0) | 0.74* | 0.77* | 0.102 |
| Print | 1 | 88.9 (73.9; 96.9) | 0.7* | 0.71* | 0.135 |
|  | 2 | 95.8 (78.9; 99.9) | 0.92* | 0.92* | 0.607 |

\* p < 0.001; 1 Stuart-Maxwell-Test

**Supplement Figure S2:**
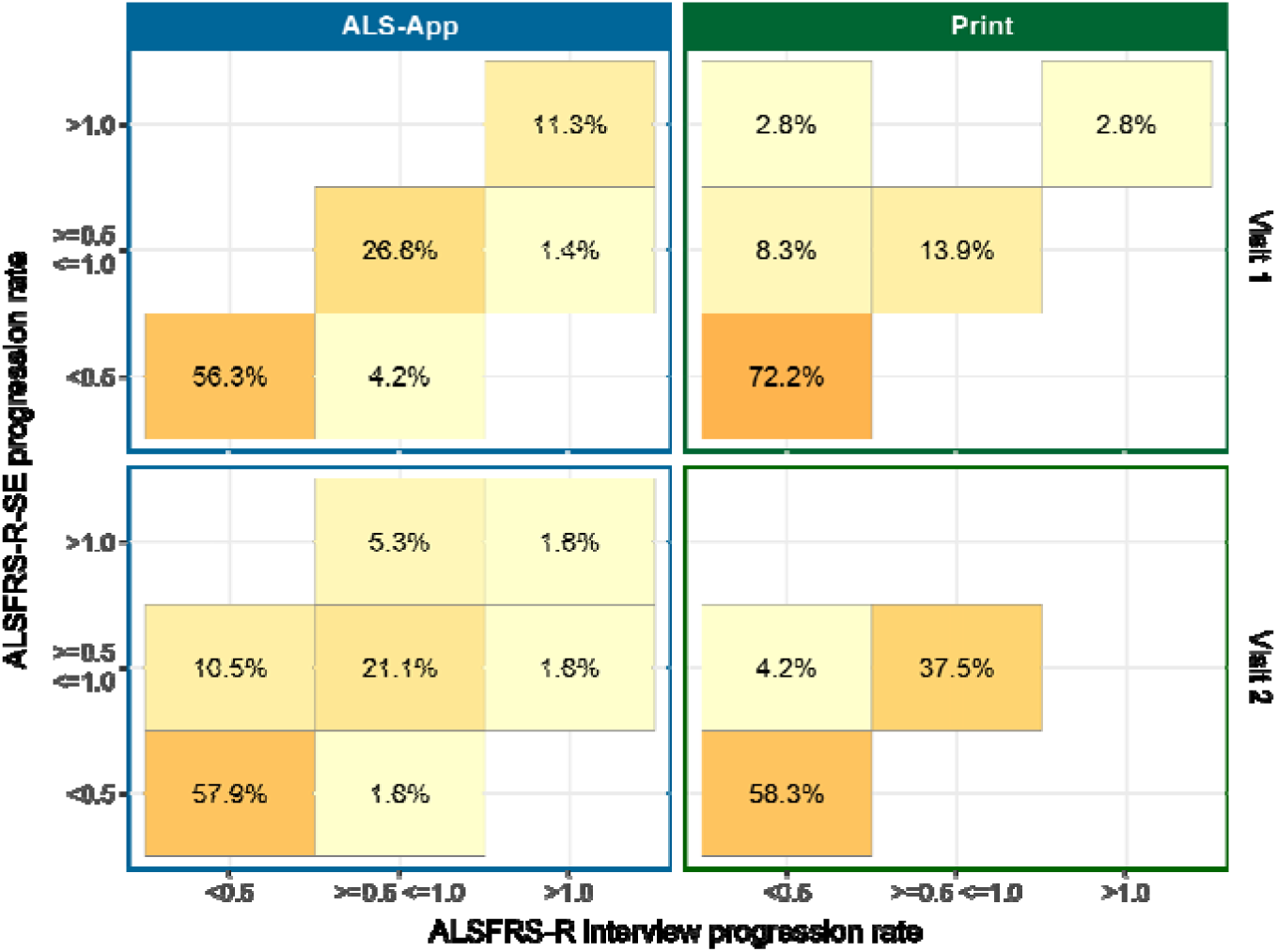
Comparison between progression rates resulting from the ALSFRS-R-SE and the harmonized ALSFRS-R interview. Progression rates were categorized as <0.5, ≥ 0.5 to ≤ 1.0, and greater than 1.0. The proportion of patients with the corresponding PR is shown in each field. A darker color indicates a higher proportion with a stronger correlation of PR, respectively. Fields on the bisecting axis indicate an exact match.

## Notes

### Funding Statement

This work was supported by the Boris Canessa ALS Stiftung (Duesseldorf, Germany) and Martin Herrenknecht Fonds for ALS Research (H4017703513237604).

### Author Declarations

Ethics committee of Committee of Charite - Universitaetsmedizin Berlin, Germany gave ethical approval for this work (EA2/190/23).

## References

1. Cedarbaum JM, Stambler N, Malta E, Fuller C, Hilt D, Thurmond B, et al. The ALSFRS-R: a revised ALS functional rating scale that incorporates assessments of respiratory function. J Neurol Sci: 1999;169:13–21. 10.1016/S0022-510X(99)00210-5.

2. Genge A, Cedarbaum JM, Shefner J, Chio A, Al-Chalabi A, Van Damme P, et al. The ALSFRS-R Summit: a global call to action on the use of the ALSFRS-R in ALS clinical trials. Amyotroph Lateral Scler Front Degener: 2024;25:382–387. 10.1080/21678421.2024.2320880.

3. Shefner JM, Bunte T, Kittle G, Genge A, Van Den Berg LH. Harmonized standard operating procedures for administering the ALS functional rating scale-revised. Amyotroph Lateral Scler Front Degener: 2024;25:26–33. 10.1080/21678421.2023.2260832.

4. Bakers JNE, De Jongh AD, Bunte TM, Kendall L, Han SS, Epstein N, et al. Using the ALSFRS-R in multicentre clinical trials for amyotrophic lateral sclerosis: potential limitations in current standard operating procedures. Amyotroph Lateral Scler Front Degener: 2022;23:500–507. 10.1080/21678421.2021.2016838.

5. Hamilton J, Mohan P, Kittle G, Shefner JM. Impact of mode of training and recertification on ALSFRS-R rater performance. Amyotroph Lateral Scler Front Degener: 2023;24:289–294. 10.1080/21678421.2022.2149344.

6. Montes J, Levy G, Albert S, Kaufmann P, Buchsbaum R, Gordon PH, et al. Development and evaluation of a self-administered version of the ALSFRS-R. Neurology: 2006;67:1294–1296. 10.1212/01.wnl.0000238505.22066.fc.

7. Maier A, Holm T, Wicks P, Steinfurth L, Linke P, Münch C, et al. Online assessment of ALS functional rating scale compares well to in-clinic evaluation: A prospective trial. Amyotroph Lateral Scler: 2012;13:210–216. 10.3109/17482968.2011.633268.

8. Bakker LA, Schröder CD, Tan HHG, Vugts SMAG, van Eijk RPA, van Es MA, et al. Development and assessment of the inter-rater and intra-rater reproducibility of a self-administration version of the ALSFRS-R. J Neurol Neurosurg Psychiatry: 2020;91:75–81. 10.1136/jnnp-2019-321138.

9. Erb MK, Calcagno N, Brown R, Burke KM, Scheier ZA, Iyer AS, et al. Longitudinal comparison of the self-administered ALSFRS-RSE and ALSFRS-R as functional outcome measures in ALS. Amyotroph Lateral Scler Front Degener: 2024;25:570–580. 10.1080/21678421.2024.2322549.

10. Maier A, Boentert M, Reilich P, Witzel S, Petri S, Großkreutz J, et al. ALSFRS-R-SE: an adapted, annotated, and self-explanatory version of the revised amyotrophic lateral sclerosis functional rating scale. Neurol Res Pract: 2022;4:60. 10.1186/s42466-022-00224-6.

11. Meyer T, Spittel S, Grehl T, Weyen U, Steinbach R, Kettemann D, et al. Remote digital assessment of amyotrophic lateral sclerosis functional rating scale – a multicenter observational study. Amyotroph Lateral Scler Front Degener: 2023;24:175–184. 10.1080/21678421.2022.2104649.

12. Steinfurth L, Grehl T, Weyen U, Kettemann D, Steinbach R, Rödiger A, et al. Self-assessment of amyotrophic lateral sclerosis functional rating scale on the patient’s smartphone proves to be non-inferior to clinic data capture. Amyotroph Lateral Scler Front Degener: February 2025:1–12. 10.1080/21678421.2025.2468404.

13. Allen MD, Van Eijk RPA, Knox L, Carlton J, Hobson E, Mcdermott CJ, et al. Variability across versions of the self-administered ALSFRS-R: a review and call for harmonization. Amyotroph Lateral Scler Front Degener: June 2025:1–6. 10.1080/21678421.2025.2522400.

14. Mokkink LB, Terwee CB, Patrick DL, Alonso J, Stratford PW, Knol DL, et al. The COSMIN checklist for assessing the methodological quality of studies on measurement properties of health status measurement instruments: an international Delphi study. Qual Life Res: 2010;19:539–549. 10.1007/s11136-010-9606-8.

15. Van Eijk RPA, De Jongh AD, Nikolakopoulos S, McDermott CJ, Eijkemans MJC, Roes KCB, et al. An old friend who has overstayed their welcome: the ALSFRS-R total score as primary endpoint for ALS clinical trials. Amyotroph Lateral Scler Front Degener: 2021;22:300–307. 10.1080/21678421.2021.1879865.

16. The Amyotrophic Lateral Sclerosis Functional Rating Scale: Assessment of Activities of Daily Living in Patients With Amyotrophic Lateral Sclerosis. Arch Neurol: 1996;53:141. 10.1001/archneur.1996.00550020045014.

17. Martin Bland J, Altman DouglasG. STATISTICAL METHODS FOR ASSESSING AGREEMENT BETWEEN TWO METHODS OF CLINICAL MEASUREMENT. The Lancet: 1986;327:307–310. 10.1016/S0140-6736(86)90837-8.

18. Lu M-J, Zhong W-H, Liu Y-X, Miao H-Z, Li Y-C, Ji M-H. Sample Size for Assessing Agreement between Two Methods of Measurement by Bland−Altman Method. Int J Biostat: 2016;12:20150039. 10.1515/ijb-2015-0039.

19. Coons SJ, Gwaltney CJ, Hays RD, Lundy JJ, Sloan JA, Revicki DA, et al. Recommendations on Evidence Needed to Support Measurement Equivalence between Electronic and Paper-Based Patient-Reported Outcome (PRO) Measures: ISPOR ePRO Good Research Practices Task Force Report. Value Health: 2009;12:419–429. 10.1111/j.1524-4733.2008.00470.x.

20. Lin LI-K. A Concordance Correlation Coefficient to Evaluate Reproducibility. Biometrics: 1989;45:255. 10.2307/2532051.

21. Payne RB. Deming’s Regression Analysis in Method Comparison Studies. Ann Clin Biochem Int J Lab Med: 1985;22:430–430. 10.1177/000456328502200419.

22. Kendall MG. THE TREATMENT OF TIES IN RANKING PROBLEMS. Biometrika: 1945;33:239–251. 10.1093/biomet/33.3.239.

23. Stuart A. A TEST FOR HOMOGENEITY OF THE MARGINAL DISTRIBUTIONS IN A TWO-WAY CLASSIFICATION. Biometrika: 1955;42:412–416. 10.1093/biomet/42.3-4.412.

24. Kimura F, Fujimura C, Ishida S, Nakajima H, Furutama D, Uehara H, et al. Progression rate of ALSFRS-R at time of diagnosis predicts survival time in ALS. Neurology: 2006;66:265–267. 10.1212/01.wnl.0000194316.91908.8a.

25. Chew S, Burke KM, Collins E, Church R, Paganoni S, Nicholson K, et al. Patient reported outcomes in ALS: characteristics of the self-entry ALS Functional Rating Scale-revised and the Activities-specific Balance Confidence Scale. Amyotroph Lateral Scler Front Degener: 2021;22:467–477. 10.1080/21678421.2021.1900259.

26. Van Eijk RPA, De Jongh AD, Nikolakopoulos S, McDermott CJ, Eijkemans MJC, Roes KCB, et al. An old friend who has overstayed their welcome: the ALSFRS-R total score as primary endpoint for ALS clinical trials. Amyotroph Lateral Scler Front Degener: 2021;22:300–307. 10.1080/21678421.2021.1879865.

27. Küffner R, Zach N, Norel R, Hawe J, Schoenfeld D, Wang L, et al. Crowdsourced analysis of clinical trial data to predict amyotrophic lateral sclerosis progression. Nat Biotechnol: 2015;33:51–57. 10.1038/nbt.3051.

28. Hobson EV, McDermott CJ. Supportive and symptomatic management of amyotrophic lateral sclerosis. Nat Rev Neurol: 2016;12:526–538. 10.1038/nrneurol.2016.111.

29. Haenel E, Elash CA, Garner K, Turner M, Kern S. Flexible approaches to eCOA administration in clinical trials: The site perspective. Contemp Clin Trials Commun: 2024;37:101241. 10.1016/j.conctc.2023.101241.

30. Möhwald LM, Maier A, Grehl T, Weyen U, Weydt P, Günther R, et al. Shared prognostic information in amyotrophic lateral sclerosis – systematic assessment of the patients’ perception of neurofilament light chain and the ALS functional rating scale. Neurol Res Pract: 2025;7:6. 10.1186/s42466-024-00363-y.

31. Meyer T, Dreger M, Grehl T, Weyen U, Kettemann D, Weydt P, et al. Serum neurofilament light chain in distinct phenotypes of amyotrophic lateral sclerosis: A longitudinal, multicenter study. Eur J Neurol: 2024;31. 10.1111/ene.16379.

32. Muehlhausen W, Byrom B, Skerritt B, McCarthy M, McDowell B, Sohn J. Standards for Instrument Migration When Implementing Paper Patient-Reported Outcome Instruments Electronically: Recommendations from a Qualitative Synthesis of Cognitive Interview and Usability Studies. Value Health: 2018;21:41–48. 10.1016/j.jval.2017.07.002.

